# Prognostic implications of structural heart disease and premature ventricular contractions in recovery of exercise

**DOI:** 10.1101/2021.12.22.21268216

**Authors:** Thomas Lindow, Magnus Ekström, Lars Brudin, Kristofer Hedman, Martin Ugander

## Abstract

**Background:** Premature ventricular contractions (PVCs) during recovery of exercise stress testing are associated with increased cardiovascular mortality, but the cause remains unknown. We aimed to evaluate the association of PVCs during recovery with echocardiographic abnormalities, and their combined prognostic performance.

**Methods:** Echocardiographic abnormalities (reduced left ventricular (LV) ejection fraction, valvular heart disease, LV dilatation, LV hypertrophy, or increased filling pressures) and PVCs during recovery (≥1/min) were identified among patients having undergone echocardiography within median [interquartile range] 0 [0-2] days of an exercise stress test. The association between such changes and cardiovascular mortality was analyzed using Cox regression adjusted for age, sex, clinical and exercise variables.

**Results:** Among included patients (n=3,106), PVCs during recovery were found in 1,327 (43%) patients, among which the prevalence of echocardiographic abnormalities was increased (58% vs. 44%, p<0.001). Overall, PVCs during recovery were associated with increased cardiovascular mortality (219 total events, 7.9 [5.4–11.1] years follow-up; adjusted hazard ratio (HR [95% confidence interval]) 1.6 [1.2–2.1], p<0.001). When combined with echocardiographic abnormalities, PVCs during recovery were associated with increased risk when such were present (HR 3.3 [1.9–5.5], p<0.001) but not when absent (HR 1.5 [0.8–2.8], p=0.22), in reference to those with neither.

**Conclusion:** PVCs during recovery were associated with increased prevalence of echocardiographic abnormalities. Increased risk of cardiovascular mortality was observed only for subjects with PVCs if concomitant echocardiographic abnormalities were present. Our findings provide mechanistic insights to the increased CV risk reported in patients with PVCs during recovery.

## Introduction

Exercise-induced premature ventricular contractions (PVCs) are common findings in exercise stress testing and have been described to associate with increased risk of cardiovascular (CV) mortality, especially when occurring during the recovery phase.^1-11^ This has been observed in patients across a wide range of clinical risk profiles, from presumably healthy, asymptomatic individuals to patients with established CV disease.^1-4, 6, 7, 11-13^ This suggests that the etiology of PVCs during recovery is multifactorial although the cause remains to be elucidated. PVCs during recovery after exercise have been suggested to be caused by imbalances in autonomic tone,^5, 6, 14^ or to be related to myocardial ischemia.^12^ Despite the extensive data on prognostic information from exercise-induced PVCs, data on structural heart disease in patients with PVCs during the recovery phase are limited and restricted to assessment of left ventricular ejection fraction (LVEF).^1, 15^ Therefore, we aimed to investigate the association of PVCs during recovery after bicycle exercise stress with abnormalities on echocardiographic examination, and to evaluate the prognostic value of the combination of PVCs during recovery and abnormal findings on echocardiography.

## Methods

We performed a retrospective cohort study including patients aged 18 years or older who had performed a clinical bicycle exercise stress test and a resting transthoracic echocardiography within 90 days of the exercise stress test at the Department of Clinical Physiology at Kalmar County Hospital, Sweden, between 31 May 2005 and 31 Oct 2016. The exercise stress test database has been described in detail elsewhere.^16-19^ We excluded those who had an exercise time less than 3 minutes or had atrial fibrillation, considering the challenge of distinguishing aberrantly conducted supraventricular complexes from PVCs. Full inclusion and exclusion criteria are shown in Fig. 1. The database has been cross-linked with comorbidity and hospital admission data in the Swedish National Patient Registry, and with the Swedish National Causes of Death Registry to obtain survival status of all patients (up to 31 Dec 2019).^20^ The underlying cause of death, e.g. CV death, can be identified in the National Causes of Death Registry and is based on the death certificate, usually completed by the patient’s regular physician or the physician who last saw the patient. A detailed description on how the underlying cause of death is identified has been described previously.^21^ In brief, it is defined as the disease (or injury) which initiated the train of events leading directly to death. For this study, any underlying cause of death within the CV disease ICD-10 chapter (I.x) has been considered as CV death. List of definitions of comorbid conditions including International Classification of Diseases ICD-10 codes can be found in Supplemental Table A.

**Figure 1.**
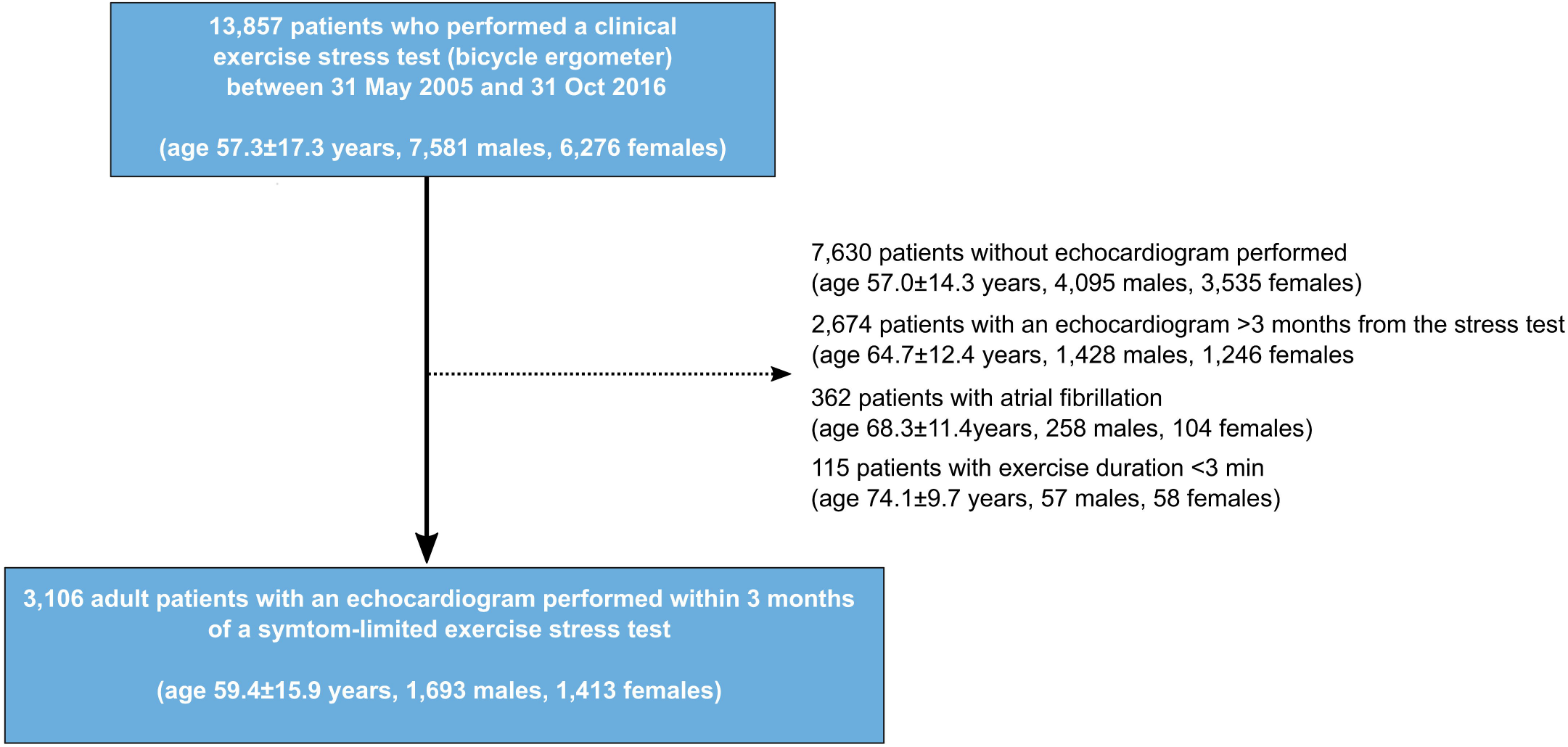
Flow chart of patient inclusion and exclusion.

### Exercise stress testing

Exercise stress testing was performed according to a nationally standardized protocol on an electronically braked, regularly calibrated bicycle ergometer (Rodby Inc, Karlskoga, Sweden) and commenced at a starting load of 40–100 W for males and 30–50W for females, with an incremental increase of 10–20 W/min.^22^ A 12-lead ECG was recorded at rest, during exercise and the recovery phase which lasted at least 4 minutes (CASE 12; Marquette Electronics Inc., Milwaukee, WI, USA and CASE v 6.51, GE Healthcare, Milwaukee, WI, USA). Every 2 minutes during exercise, the patients were queried regarding rating of perceived exertion (RPE), and systolic blood pressure was measured. The test was terminated at the patient’s will or if any termination criteria were fulfilled (severe chest pain, ST-depression ≥0.4 mV, a drop in systolic blood pressure or occurrence of severe arrhythmia (ventricular tachycardia [VT] defined as at least 3 PVCs in a row, PVCs in increasing frequency or complexity, supraventricular tachycardia >200/min, or AV block II or III occurring during exercise).^22^ At termination of exercise, the patient directly returned to supine position.

ECG abnormalities at rest before, after, and during stress were noted by the attending physician at the time of the test and were not re-interpreted for the purpose of this study. Peak workload was related to nationally standardized reference values (% of predicted W_max_).^19^ The amplitude and slope of the ST segment during the exercise stress test were measured 60 ms following the J-point (ST60). For this study, ST depression was defined as horizontal or down-sloping ST depression ≥0.1 mV in V5 during exercise or during the recovery phase. Heart rate recovery was defined as the difference in heart rate between the maximal heart rate and the heart rate 1 min after cessation of exercise.^23, 24^ For patients who had performed more than one test, only the most recent test was included.

### Frequency of PVCs in the recovery phase

PVC frequency during recovery was graded at the time of the stress test according to clinical routine as either <1 PVC/min, 1–5 PVCs/min, 5–10/min, or >10/min during the first 4 minutes of recovery. Studies have applied different definitions of frequent PVCs,^1-7, 9, 10^ ranging from the use of the median frequency in the dataset (frequent = above the median),^1, 6, 10^ to 10% of all depolarizations within a specific time frame,^4, 7, 9^ or a fixed value, e.g. >10/min ^3^. Thus, any cutoff would be arbitrary and previous studies have shown that cardiovascular risk may be similar for patients with frequent PVCs compared to those with infrequent PVCs ^6^. This suggests that the prognostic significance lies in the presence of PVCs during recovery rather than the quantity. Consequently, we chose to primarily divide our study population into two groups based on <1 or ≥1 PVC/min during the first 4 minutes of recovery. For readability, these patients will subsequently be referred to as having no PVCs during recovery, although they, by definition, may have had 0 – 3 PVCs during the first 4 minutes of recovery. As a supplemental analysis, we also present the results based on the grading used in the clinical reports, i.e., <1 PVC/min, 1–5 PVCs/min, 5–10/min, or >10/min.

### Echocardiography

echocardiographic examinations were acquired during clinical routine by skilled ultrasound technicians under physician supervision. LV ejection fraction (EF) was reported either based on the Simpson biplane method, from M-mode data, or by visual estimation. Data on grade of aortic, mitral or tricuspid regurgitation (0 – 3: none, mild, moderate, severe) were extracted from the echocardiographic reports. Moderate aortic stenosis was defined as aortic valve (AV) maximal velocity ≥3.1–4.0 m/s or AV mean gradient 20–40 mm Hg, and severe aortic stenosis as AV maximal velocity ≥4.0 m/s or AV mean gradient ≥40 mm Hg.^25^

LV mass was calculated according to the Cube formula (LV mass (g) = 0.8 × 1.04 ([IVS + LVID + PW]^3^ – LVID^3^) + 0.6).^26^ Left-sided cardiac dimensions (LV diameter, LA diameter, LV mass) were indexed to body-surface area (BSA) and were considered abnormal if they exceeded established sex-based reference values (LV diameter: ≥31 mm/m^2^ (males), ≥32 mm/m^2^ (females); LA diameter: ≥23 mm/m^2^ (both sexes); LV mass: >115 g/m^2^ (males), >95 g/m^2^ (females)) ^26^. Similarly, LVEF below the sex-based reference value was considered to be reduced (<52% (males), <54% (females)).^26^

Significant valvular heart disease was defined as at least moderate aortic/mitral/tricuspid regurgitation or at least moderate aortic stenosis. Increased right ventricle to right atrial (RV– RA) pressure gradient was defined as a tricuspid regurgitant velocity exceeding 2.8 m/s. Increased LV filling pressures were defined as E/e’ ≥15 and a moderately enlarged LA, or E/e’ ≥15 and an increased RV–RA pressure gradient. An echocardiographic abnormality was considered present if any of the following criteria were met: a) reduced LVEF, b) significant valvular heart disease, c) LV dilatation, d) increased LV filling pressures or e) LV hypertrophy. Reasons for referral/primary referral questions are presented as supplemental material.

The study complies with the Declaration of Helsinki and was approved by the Regional Ethical Review Board (Dnr 2012/379-31; 2018/141-31 and 2020/00352). Informed consent was waived by the Ethical Review Board.

### Statistical analysis

Continuous variables were described as mean±standard deviation [SD], or as median [interquartile range]. Proportional differences between groups were assessed using the χ2 test. Comparison of group means was performed using Student’s t test or analysis of variance (ANOVA). In order to account for baseline differences in age and sex, the relation between the frequency of PVCs in the stress test recovery phase and subsequent findings on echocardiography was described not only as proportions, but also using odds ratios (OR) with 95% confidence intervals (CI) from multivariable logistic regression.

Time-to-event analysis was performed using Kaplan-Meier analysis with censoring at study end (31 Dec 2019). The association between PVCs during recovery and CV mortality was analyzed using multivariable Cox proportional hazard regression models; unadjusted and adjusted for age, sex, hypertension, heart failure, ischemic heart disease, diabetes mellitus, body mass index, peak workload, maximal heart rate, maximal systolic blood pressure, heart rate recovery, ST depression, cardiovascular medications, and echocardiographic abnormalities. Further, the combination of PVCs during recovery and presence/absence of echocardiographic abnormalities were analyzed unadjusted and adjusted for age, sex, baseline comorbidities (hypertension, heart failure, ischemic heart disease, diabetes mellitus, body mass index), peak workload, maximal heart rate, maximal systolic blood pressure, heart rate recovery, and ST depression, and cardiovascular medications. The choice of confounding variables was based on subject matter knowledge and directed acyclic graphs.

In addition, instead of analyzing the association between any echocardiographic abnormalities as a group, single predictors (increased E/e ratio, LV dilatation, increased LV mass, increased LA diameter, reduced LVEF, increased RV-RA pressure gradient, aortic regurgitation, aortic stenosis, mitral regurgitation, tricuspid regurgitation) were also evaluated in their relation to CV death in patients with PVCs during recovery, both in univariate and multivariable analysis. Furthermore, the combination of PVCs during recovery and echocardiographic abnormalities were separately evaluated for its association with incident acute coronary syndrome (ACS; acute myocardial infarction or unstable angina) during follow-up and within one year. Hazard ratios (HR) with 95% CI were presented. The assumption of proportional hazards was confirmed using Schoenfeld’s residuals. The following sensitivity analyses were performed in relation to the Cox regression analysis: 1) excluding patients with PVCs at rest according to the clinical resting ECG report, since PVCs during the recovery phase could reflect PVCs at rest although unrelated to exercise, and 2) including only patients who underwent echocardiography more than 3 months after the exercise stress test, since there may be a selection bias when only those that were referred for both exercise stress test and echocardiography at almost the same time were included, and 3) stratifying patients based on the grading of PVC frequency used in the clinical reports (<1 PVC/min, 1–5/min, ≥5–10, >10/min), and 4) stratifying the analysis by recency (echocardiogram performed before or after the year 2010), since technical advances may have had an impact of the results. The results of the sensitivity analyses are presented as supplemental material. We also reported the results for all-cause mortality since this outcome is likely to be less dependent on the consistency in diagnostic reporting.

The likelihood of PVCs during recovery was assessed by univariate and multivariable logistic regression analysis. Age, male sex, relevant comorbidities (diabetes, hypertension, ischemic heart disease), echocardiographic variables (at least moderate valvular heart disease, LVEF, E/e’, left atrial diameter, left ventricular diameter) and exercise variables assumed to be related to parasympathetic tone (resting heart rate, difference in heart rate from supine rest to sitting, heart rate recovery, % of predicted maximal heart rate) were included in univariate analysis and subsequently in the multivariable analysis if the p value was less than 0.1.

Statistical significance was accepted at the level of p<0.05 (two-sided). Statistical analysis was performed using R v. 3.5.3 (R Core Team (2021). R: A language and environment for statistical computing. R Foundation for Statistical Computing, Vienna, Austria (https://www.R-project.org/).

## Results

In total, 3,106 patients were included (age 59±16 years, 54.5% males) and followed for a median 7.9 years [IQR 5.4–11.1, range 0 – 15], range 0.0–14.6 years.

PVCs during the recovery phase were present in 1,327 (42.7%) patients. During follow-up, 219 (7.1%) patients died from a CV cause (10.4% of patients with vs. 4.6% without PVCs during recovery phase, p<0.001). Baseline characteristics based on the presence of PVCs during the recovery phase are presented in Table 1. Besides older age and a larger proportion of male subjects among those with than without PVCs during recovery, there were no major differences in comorbidities between groups. Peak workload was lower in patients with PVCs, but the difference in mean peak workload as % of predicted was not clinically relevant. A larger proportion of tests were terminated by the physician due to arrhythmia among those with PVCs during recovery.

**Table 1.**
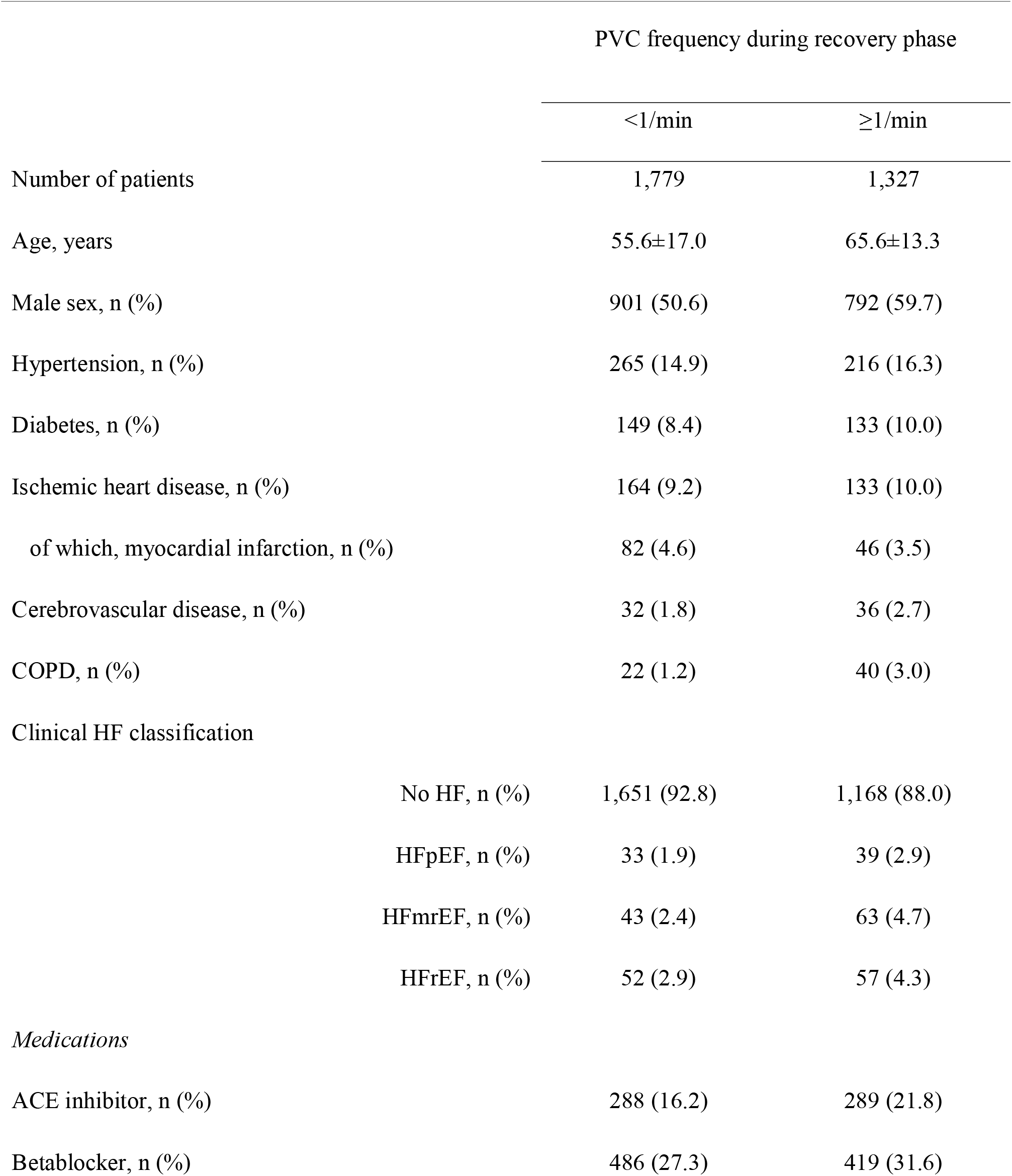
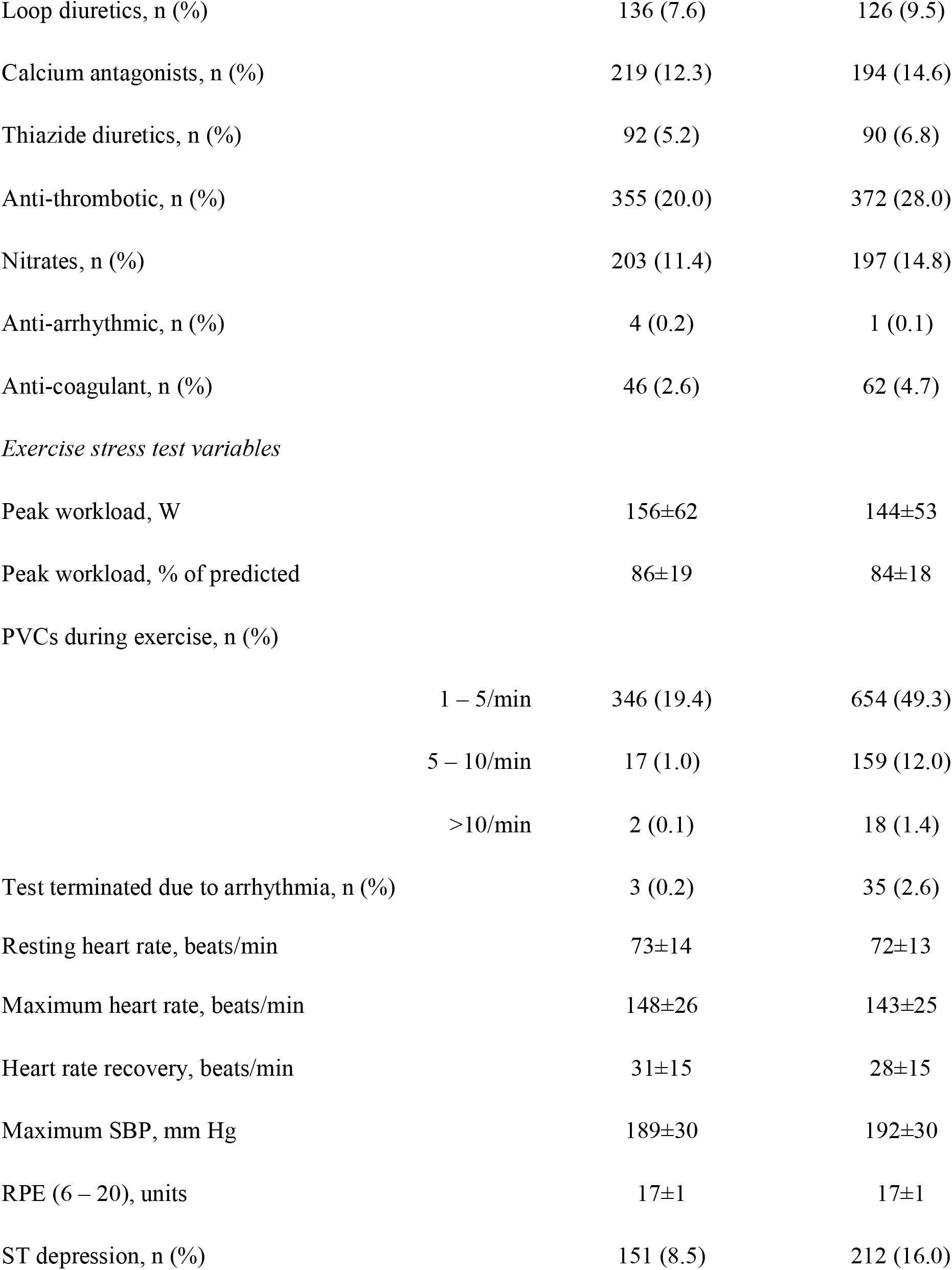
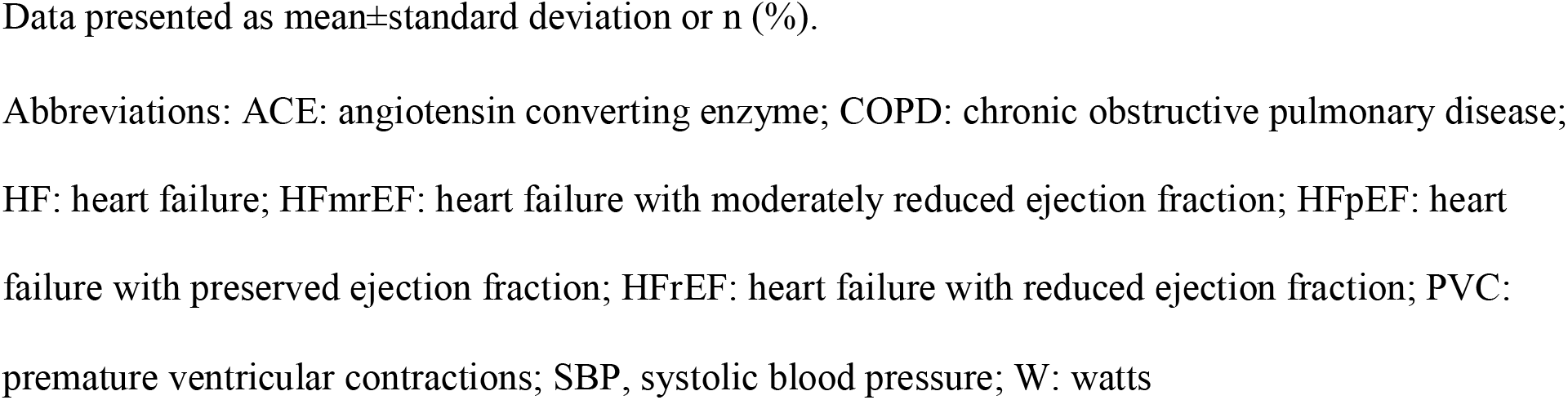
Baseline characteristics including exercise stress test characteristics stratified by frequency of premature ventricular contractions during the recovery phase

An echocardiographic examination was performed within 1 day of the exercise stress test in 74.6% of patients, median [interquartile range] 0 [0-2] days. An increased prevalence of echocardiographic abnormalities was found for patients with PVCs during the recovery phase (57.6% vs 42.9%, p<0.001 (Table 2). Mean LVEF was only slightly lower in patients with PVCs during the recovery phase (62% vs 64%, p<0.001), but the proportion of patients with reduced LVEF was higher in patients with PVCs during recovery compared to those without PVCs (14% vs. 9%, p<0.001). PVCs during recovery was associated with increased odds of having echocardiographic abnormalities, even after adjustment for age and sex (OR 1.4 [1.2– 1.6], p<0.001).

**Table 2.**
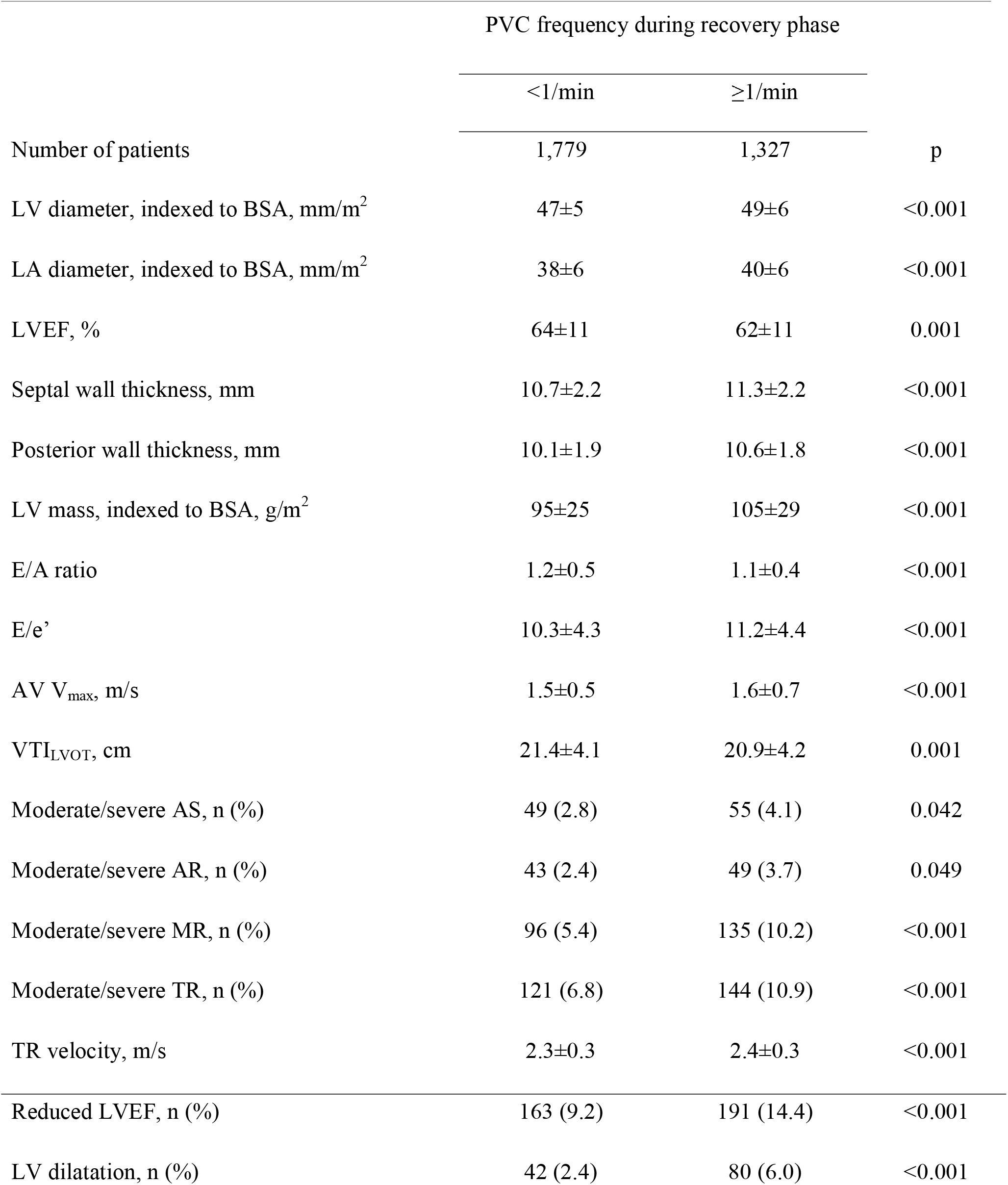
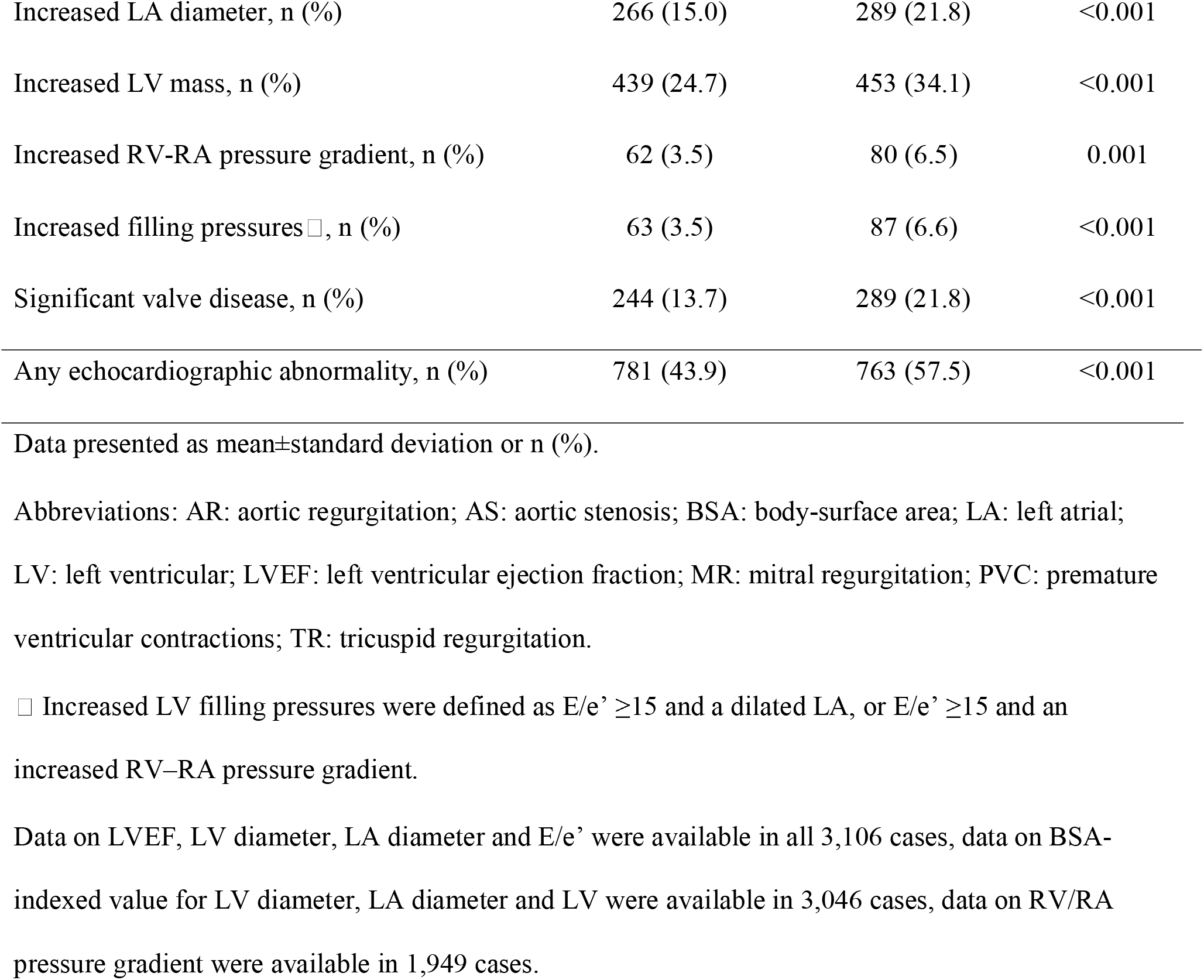
Echocardiographic outcomes stratified by occurrence of premature ventricular contractions during the recovery phase.

PVCs during recovery were associated with increased CV mortality (unadjusted HR 2.6 [2.0– 3.4]), also after adjustment for age, sex, clinical variables, exercise stress testing variables and echocardiographic abnormalities (1.6 [1.2–2.1]). However, when analyzed in combination with either presence or absence of echocardiographic abnormalities, PVCs during recovery were only associated with increased risk of CV death when echocardiographic abnormalities were present (Table 3, Fig. 2 and 3). The risk for patients with PVCs during recovery but no echocardiographic abnormalities was not increased relative to those with <1 PVC/min PVCs during recovery and absent echocardiographic abnormalities. The proportion of echocardiographic abnormalities in patients with PVCs during recovery stratified by CV death during follow-up is presented in Figure 4. Among those with PVCs during recovery who died from a CV cause, the most common echocardiographic abnormality was increased LV mass, followed by LA dilatation, elevated E/e’, any valvular heart disease, and reduced LVEF. In patients with PVC during recovery, increased LV mass, increased E/e’ ratio, LV dilatation were associated with CV mortality in the multivariable analysis, whereas LVEF, LA dilatation, increased RV/RA gradient and valvular heart disease were not (Table 4).

**Table 3.** Hazard ratios [95%CI] for cardiovascular mortality based on combination of absence/presence of PVCs during recovery, and presence or absence of significant echocardiographic abnormalities, n=3,106 (219 events)

|  | Unadjusted | Adjusted for age, sex, clinical<sup>†</sup> and exercise variables<sup>§</sup> |
| --- | --- | --- |
| PVC-/ECHO- | 1.0 | 1.0 |
| PVC+/ECHO- | 2.8 [1.5–5.2] | 1.5 [0.8–2.8] |
| PVC-/ECHO+ | 4.4 [2.5–7.4] | 2.0 [1.2–3.4] |
| PVC+/ECHO+ | 9.1 [5.5–15.2] | 3.3 [1.9–5.5] |

Abbreviations: ECHO-: no significant abnormality on echocardiography; ECHO+: significant abnormality on echocardiography; PVC: premature ventricular contractions; PVC-: <1 PVCs/min during recovery; PVC+: ≥1 PVCs/min during recovery.
<sup>†</sup> Hypertension, diabetes, ischemic heart disease, heart failure, body-mass index
<sup>§</sup> Peak workload, maximal heart rate, heart rate recovery, maximal systolic blood pressure, ST depression

**Table 4.** Hazard ratio [95% condifence interval] for cardiovascular mortality based on presence or absence of significant echocardiographic abnormalities in patients with PVCs during recovery, n=1,327 (138 events)

|  | Unadjusted<br>(univariate) | Adjusted for age, sex and<br>baseline comorbidities<sup>†</sup> |
| --- | --- | --- |
| Increased E/e' ratio | 3.7 [2.6–5.3] | 2.1 [1.4–3.1] |
| LV dilatation | 3.6 [2.3–5.6] | 2.1 [1.2–3.6] |
| Increased LV mass | 2.6 [1.8–3.6] | 1.7 [1.2–2.4] |
| Increased LA diameter | 2.6 [1.8–3.6] | 1.0 [0.7–1.6] |
| Reduced LVEF | 3.3 [2.2–4.8] | 1.5 [1.0–2.5] |
| Increased RV/RA gradient | 3.4 [2.2–5.3] | 1.4 [0.9–2.6] |
| Aortic regurgitation | 2.4 [1.4–4.4] | 1.4 [0.8–2.6] |
| Mitral regurgitation | 2.6 [1.7–3.8] | 1.1 [0.7–1.9] |
| Tricuspid regurgitation | 1.7 [1.1–2.7] | 0.9 [0.5–1.5] |
| Aortic stenosis | 1.1 [0.5–2.4] | 0.7 [0.3–1.5] |

Abbreviations: LA: left atrial; LV: left ventricle/ventricular; LVEF: LV ejection fraction.
<sup>†</sup> Hypertension, diabetes, ischemic heart disease, heart failure, body-mass index

**Figure 2.**
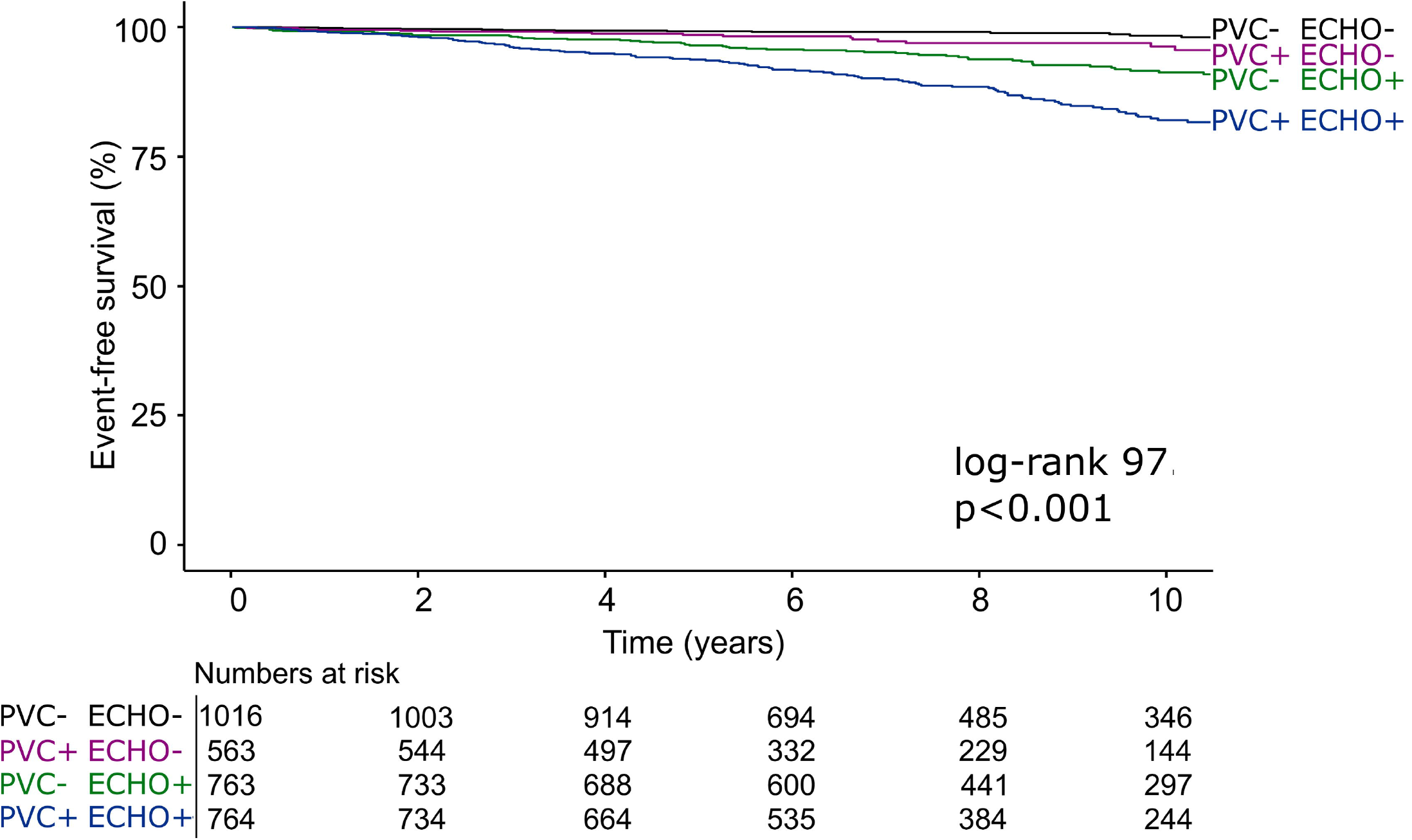
Time-to-event analysis for the combination of PVCs during recovery and abnormalities on echocardiography among 3,106 patients experiencing 219 cardiovascular mortality events during 7.9 [5.4–11.1] years of follow-up. Abbreviations: ECHO-: no significant abnormality on echocardiography; ECHO+: significant abnormality on echocardiography; PVC: premature ventricular contractions; PVC-: <1 PVC/min during recovery; PVC+: ≥1 PVC/min during recovery. An echocardiographic abnormality was defined as either: reduced left ventricular ejection fraction, at least moderate valvular heart disease, left ventricular dilatation, increased left ventricular mass, or increased left ventricular filling pressures.

**Figure 3.**
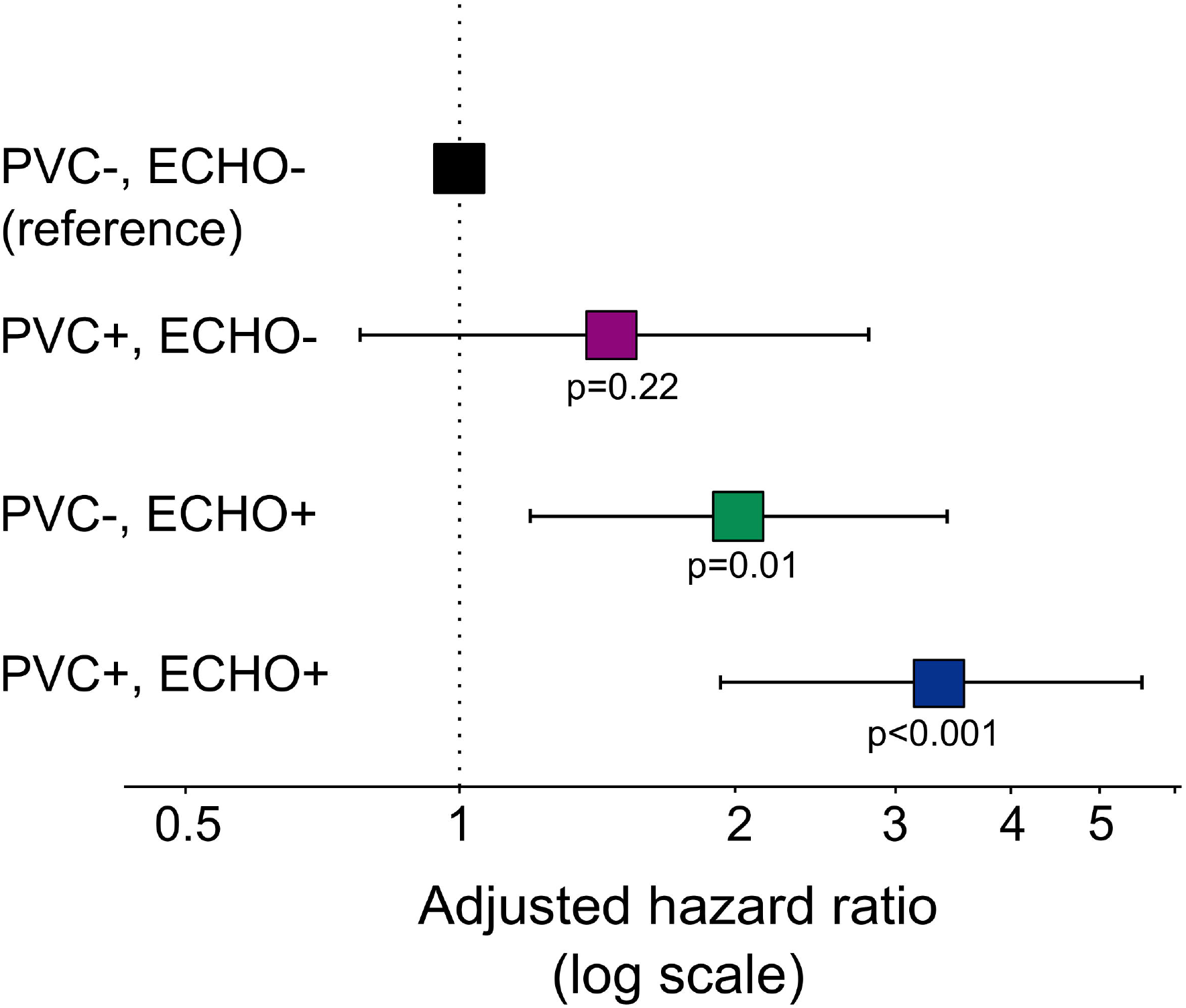
Forest plot showing hazard ratios for cardiovascular death with 95% confidence limits (adjusted for age, sex, hypertension, heart failure, ischemic heart disease, diabetes mellitus, body mass index, peak workload, maximal heart rate, maximal systolic blood pressure, heart rate recovery and ST depression) based on combinations of presence/absence of PVC during recovery and echocardiographic abnormalities. Abbreviations: ECHO-: no significant abnormality on echocardiography; ECHO+: significant abnormality on echocardiography; PVC: premature ventricular contractions; PVC-: <1 PVC/min during recovery; PVC+: ≥1 PVC/min during recovery. An echocardiographic abnormality was defined as either: reduced left ventricular ejection fraction, at least moderate valvular heart disease, left ventricular dilatation, increased left ventricular mass or increased filling pressures defined as E/e’ ≥15 and either moderately enlarged LA or increased RV–RA pressure gradient.

**Figure 4.**
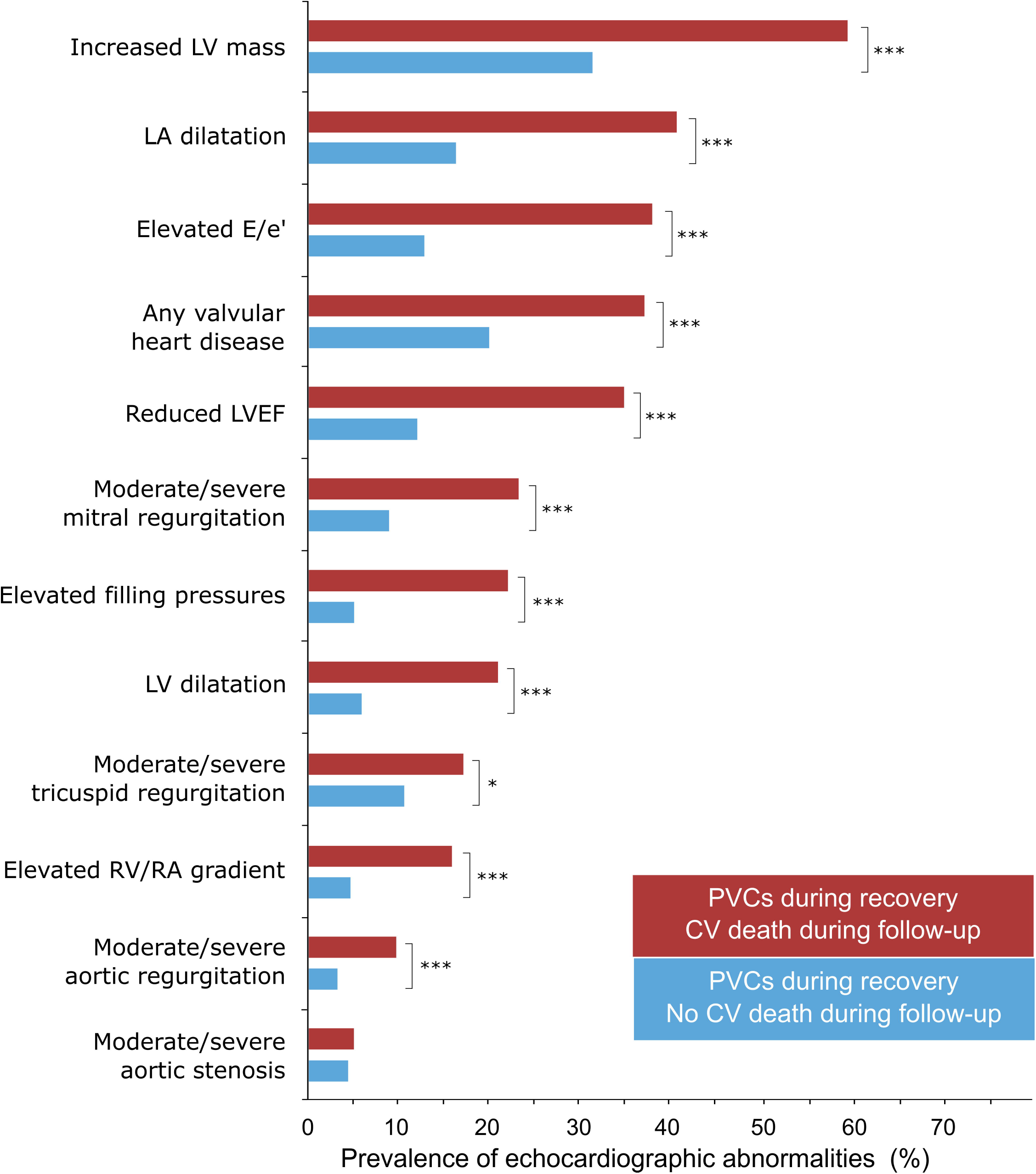
Prevalence of echocardiographic abnormalities in patients with PVC during recovery (n=1,327) stratified by cardiovascular (CV) death during follow-up (CV death: n=138 (red bars); no death: 1,189 (light blue bars). *** = <0.001, * = <0.05 Abbreviations: LV: left ventricular; LVEF: left ventricular ejection fraction; LA: left atrial; RV: right ventricle; RA: right atrium. Echocardiographic abnormalities were more common in patients with PVC during recovery and an adverse outcome.

In multivariable analysis, age, male sex, LV diameter, LV mass, and maximal heart rate (in % of predicted) were the only independent predictors of PVCs during the recovery phase. Beyond maximal heart rate, indicators of abnormal autonomic tone, such as heart rate recovery, were not associated with increased frequency of PVCs during the recovery phase in multivariable analysis (Supplemental Table B).

During follow-up, 285 patients (9.2%) were diagnosed with acute coronary syndrome (10.6% of patients with PVCs during recovery vs. 8.2% of patients without PVCs during recovery, p=0.03). In patients with no prior diagnosis of ACS (n=2,954), PVCs during recovery was associated with increased risk of incident ACS both in the absence of concomitant echocardiographic abnormalities (HR 1.7 [1.1–2.6]), and in combination with echocardiographic abnormalities (HR 2.9 [2.0–4.2]). However, for ACS within 1 year, PVCs during recovery were only associated with increased risk in the presence of concomitant echocardiographic abnormalities (HR 3.3 [1.4 – 8.0]), but not for those with PVCs without echocardiographic abnormalities (HR 0.8 [0.2 – 3.0]), in reference to patients with neither.

## Discussion

The main finding of the current study was that exercise-induced PVCs during the recovery phase were associated with increased CV mortality, and that this was only apparent when present together with echocardiographic abnormalities. These findings have two important implications. Firstly, they provide insights which may aid in the understanding of the well-known increased long-term CV risk in patients with exercise-induced PVCs during recovery. Secondly, the occurrence of PVCs during recovery implies a high diagnostic yield of subsequent echocardiography, therefore warranting further evaluation in search of structural heart disease.

Although patients with PVCs during the recovery phase had a negligible difference in mean LVEF compared to those without PVCs, the proportion of patients with reduced LVEF was markedly higher. Previous reports from cardiac imaging in the setting of exercise-induced PVCs have been restricted to the assessment of LVEF ^1^. Frolkis, *et al*, studied the prognostic value of PVCs during the recovery phase in almost 30,000 patients without a history of heart failure, valvular heart disease, or arrhythmia, and found it to be a stronger predictor of all-cause death than PVCs during exercise. In 6,421 of these patients, LVEF was assessed either by echocardiography or by contrast ventriculography within 3 months following the stress test. That study had similar findings as the current study, albeit with a higher prevalence of reduced LVEF than in the current study, both among patients with PVCs during recovery and in those without (28% *vs*. 13%). Our study provides additional insights that echocardiographic findings beyond LVEF are associated – even more strongly so than LVEF – with a worsened prognosis in patients with PVCs during the recovery phase.^1-6^ This study adds to the current literature by showing that PVCs during recovery are associated with a higher rate of structural heart disease including LV hypertrophy, LV dilatation, increased LV filling pressures, and valvular heart disease. Moreover, several of these findings were more common in those with PVCs during recovery who died during follow-up, compared to those who survived, increased LV mass, elevated E/e’ ratio and LV dilatation, in particular The possibility that the increased risk among those with PVCs is mediated by LV hypertrophy and/or mechanical stress is theoretically appealing since structural and electrical cardiac remodeling are known to be closely related.^27, 28^ LV hypertrophy has been described to increase repolarization heterogeneity, to disturb calcium ion concentration, and to cause both ion channel and gap junction remodeling, leading to increased susceptibility to develop ventricular arrythmias.^27, 29^ Also, this is in agreement with previous findings of increased CV risk among patients with PVCs at rest in combination with structural heart disease.^30-32^ From our results, it is not possible to conclude whether it is the increased risk of arrythmia associated with structural heart disease, or if PVCs is a marker of significant structural heart disease, that would explain the increased CV risk.

Interestingly, we found no association between exercise variables assumed to be related to autonomic tone and the occurrence of PVCs during the recovery phase, after adjusting for age, sex and echocardiographic findings. Autonomic abnormalities have been suggested as a plausible cause of the increased CV mortality that has been observed both in low- and high-risk patients.^5, 6, 14^ However, the results of the current study do not support such an association.

### Limitations

The findings of the current study are based on data from patients who performed both an exercise stress test and an echocardiogram based on clinical referral. This may induce selection bias, and groups of patients that are not referred for an echocardiogram may have a different proportion of abnormal findings. However, in a sensitivity analysis for those who performed an echocardiogram >90 days after the exercise test, results were highly similar regarding the prognostic value (Supplemental Table D), which strengthens the generalizability of our results. A detailed description of the clinical and exercise characteristics in patients who did not perform any echocardiogram can be found in Supplemental Table F.

Furthermore, we lack information on coronary anatomy as well as future investigations for coronary artery disease. Instead, incident ACS diagnoses were explored. We also lack information on differentiated CV causes of death, e.g. sudden cardiac death. However, very few patients underwent cardiac resuscitation and/or defibrillation during follow-up (11 out of 3,106).

The use of registry data for cause of death is highly dependent on the quality of the records. At chapter level, as applied in this study, agreement between hospital records and registry data is high^21^. Also, similar results were obtained when using all-cause mortality as an outcome.

This is a registry-based study including several thousands of patients and for practical reasons relies on clinical reports with tabulated values, instead of raw ECG or imaging data.. We have no information on the absolute number of PVCs but relied on the grading in the clinical report (<1/min, 1 - 5/min, 5 - 10/min, ≥10/min) and patients referred to in text as having no PVCs (≤1/min) may thus have had up to 3 PVCs during the first 4 minutes of recovery. We believe that equating 0 – 3 to none, in general, would be in agreement with any clinical interpretation made during test result interpretation.

Further, the classifications used relied on the number of PVCs only, i.e. not taking into account the complexity of ventricular arrythmia such as couplets, bigeminy or VT. The number of tests prematurely terminated due to severe arrhythmia among those with no or low-grade PVCs was small, indicating that this likely did not have had a substantial impact on the findings. Furthermore, measurements of RV and RA dimensions were not available and rare diagnoses such as arrhythmogenic right ventricular cardiomyopathy were therefore not evaluated. Nonetheless, they are expected to constitute a very small number of patients ^33^. Despite the above-mentioned limitations, this large registry is unique in combining results of exercise stress testing and echocardiography, and although causal inferences cannot be made, we consider our findings to provide important, new insights to the previously established increased long-term CV risk reported in patients with PVCs during recovery.

## Conclusion

PVCs during the recovery phase of exercise stress testing were associated with increased risk of abnormal findings on echocardiography. Importantly, increased CV mortality was observed only for subjects with PVCs who had concomitant echocardiographic abnormalities. Our findings provide mechanistic insights to the increased CV risk reported in patients with PVCs during recovery.

## Data Availability

All data produced in the present study are available upon reasonable request to the authors

