## Supplemental tables for "Prognostic implications of structural heart disease and premature ventricular contractions in recovery of exercise"

**SUPPLEMENTS**

**Supplemental Table A.** How diagnoses included for the purposes of demographic characteristics were defined.

| Diagnosis | Definition (either of) |
| --- | --- |
| Hypertension | 1. Prevalent diagnosis, hospital (outpatient, inpatient, admission; ICD10-codes) Hypertensive diseases (I10x-I15x) |
| Hyperlipidemia | Prevalent diagnosis, hospital (outpatient, inpatient, admission; ICD10-codes) Disorders of lipoprotein metabolism and other lipidemias (E78x) |
| Diabetes mellitus | 1. Prevalent diagnosis, hospital (outpatient, inpatient, admission; ICD10-codes) Type I (E10x), Type II (E12X), Other (E13x-E14x) |
| Heart failure | 1. Prevalent diagnosis, hospital (outpatient, inpatient, admission; ICD10-codes) Heart failure (I50x)  2. Prevalent diagnosis, hospital (outpatient, inpatient, admission; ICD10-codes) Cardiomyopathy (I42x-I43x)  3. Left ventricular ejection fraction <50% at the echocardiographic examination  4. Incident heart failure diagnosis within 3 months of the echocardiographic study (I50x) |
| Chronic obstructive pulmonary disease | 1. Prevalent diagnosis, hospital (outpatient, inpatient, admission; ICD10-codes) Bronchitis (J40x-J42x); Emphysema (J43); Other COPD (J44x) |
| Atrial fibrillation/flutter | 1. Prevalent diagnosis, hospital (outpatient, inpatient, admission; ICD10-codes) Atrial fibrillation and flutter (I48x) |
| Ischemic heart disease | 1. Prevalent diagnosis, hospital (outpatient, inpatient, admission; ICD10-codes) Angina pectoris (I20.9).  2. Prevalent diagnosis, hospital (outpatient, inpatient, admission; ICD10-codes) Acute myocardial infarction (I21x); Subsequent myocardial infarction (I22x); Complication following acute myocardial infarction (I23x); Other acute ischemic heart disease (I24x)  3. Prevalent diagnosis, hospital (outpatient, inpatient, admission; ICD10-codes) Unstable angina (I20.0) |
| Acute myocardial infarction | 1. Prevalent diagnosis, hospital (outpatient, inpatient, admission; ICD10-codes) Acute myocardial infarction (I21x); Subsequent myocardial infarction (I22x); Complication following acute myocardial infarction (I23x); Other acute ischemic heart disease (I24x) |
| Cerebrovascular disease | 1. Prevalent diagnosis, hospital (outpatient, inpatient, admission; ICD10-codes) (ICD>="I60") & (ICD<="I69") Subarachnoid haemorrhage (I60x); Intracerebral haemorrhage (I61x); Other nontraumatic intracranial haemorrhage (I62x); Cerebral infarction (I63x); Stroke, not 3 specified (I64x); Occlusion and stenosis of arteries, not resulting in cerebral infarction (I65x-166x); Other cerebrovascular diseases (I67x); Cerebrovascular disorders in diseases classified elsewhere (I68x); Sequelae of cerebrovascular disease (I69x). |
| Cardiovascular disease | 1. Prevalent diagnosis, hospital (outpatient, inpatient, admission; ICD10-codes)  ICD Ix |

| **Supplemental Table B.** Univariate and multivariable analysis for the prediction of frequent PVCs during the recovery phase of exercise stress testing | | | | | | | | |
| --- | --- | --- | --- | --- | --- | --- | --- | --- |
|  |  | | | Multivariable analysis | | | | |
|  | | Univariate analysis | | Model 1 | | Final model | | |
| Parameter | Odds ratio  [95% CI] | | p | Odds ratio  [95% CI] | p | | Odds ratio  [95% CI] | p |
| Age, years | 1.04 [1.04–1.05] | | <0.001 | 1.04 [1.03–1.05] | <0.001 | | 1.04 [1.04–1.05] | <0.001 |
| Male sex | 1.44 [1.25–1.67] | | <0.001 | 1.18 [0.99–1.40] | 0.06 | | 1.24 [1.04–1.45] | <0.001 |
| Valvular heart disease | 1.75 [1.45–2.11] | | <0.001 | 1.42 [0.98–1.47] | 0.08 | | 1.20 [0.98–1.47] | 0.08 |
| LV mass indexed to BSA, g/m^2^ | 1.01 [1.01–1.02] | | <0.001 | 1.00 [1.00–1.01] | 0.05 | | 1.00 [1.00–1.01] | 0.04 |
| LV diameter, mm | 1.04 [1.03–1.06] | | <0.001 | 1.05 [1.03–1.07] | <0.001 | | 1.06 [1.04–1.08] | <0.001 |
| LA diameter, mm | 1.06 [1.05–1.07] | | <0.001 | 1.01 [0.99–1.02] | 0.23 | |  |  |
| LVEF, % | 0.98 [0.98–0.99] | | <0.001 | 0.99 [0.98–1.00] | 0.53 | |  |  |
| % of pred. max heart rate | 1.01 [100–1.02] | | <0.001 | 1.02 1.02–1.03] |  | | 1.02 [1.01 – 1.03] | <0.001 |
| E/e’ | 1.04 [1.03–1.06] | | <0.001 | 0.98 [0.96–1.01] | 0.13 | |  |  |
| Heart rate recovery (min^-1^) | 0.98 [0.98–0.99] | | <0.001 | 1.00 [0.99–1.00] | 0.17 | |  |  |
| Resting heart rate (min^-1^) | 0.99 [0.99–1.00] | | 0.75 |  |  | |  |  |
| Heart rate increase to sitting (min^-1^) | 1.00 [0.99–1.00] | | 0.24 |  |  | |  |  |
| Ischemic heart disease | 1.11 [0.87–1.40] | | 0.41 |  |  | |  |  |
| Diabetes | 1.22 [0.95–1.56] | | 0.11 |  |  | |  |  |
| Hypertension | 1.11 [0.91–1.35] | | 0.29 |  |  | |  |  |
| Abbreviations: BSA: body-surface area; CI: confidence interval; LVEF: left ventricular ejection fraction; pred.: predicted.  Variables with a p<0.1 in the univariate analysis were included in the multivariable analysis, and subsequently included in the final prediction model if p<0.1 at multivariable analysis. | | | | | | | | |

| **Table C. Sensitivity analysis – PVCs at rest**  Hazard ratios with 95% confidence intervals from Cox regression analysis, for the combination of PVCs during recovery and echocardiographic abnormalities in patients without PVCs at rest before exercise according to the clinical report (n= 3,038, 215 events). | | |
| --- | --- | --- |
|  | Unadjusted | Adjusted for age, sex, clinical and exercise variables^†^ |
| PVC-/ECHO- | 1.0 | 1.0 |
| PVC/+/ECHO- | 2.3 [1.3–4.3] | 1.5 [0.8–2.7] |
| PVC-/ECHO+ | 4.0 [2.4–6.7] | 2.0 [1.2–3.4] |
| PVC+/ECHO+ | 8.5 [5.3–13.8] | 3.3 [2.0–5.3] |
| Abbreviations: ECHO-: no significant abnormality on echocardiography; ECHO+: significant abnormality on echocardiography; PVC-: <1 PVCs/min during recovery; PVC+: ≥1 PVCs/min during recovery  ^†^Hypertension, diabetes, ischemic heart disease, heart failure, body-mass index, peak workload, maximal heart rate, heart rate recovery, ST depression, use of either betablocker, ace inhibitor, angiotension blockers, loop diuretics, antihrombotics, anti-coagulants, or calcium channel blockers. | | |

| **Table D. Sensitivity analysis – echocardiography performed unrelated to the exercise stress test**  Hazard ratios with 95% confidence intervals from Cox regression analysis, for the combination of PVCs during recovery and echocardiographic abnormalities in patients (n= 2,624, 228 events) who performed an echocardiographic study more than 3 months after the stress test (median 757 days, interquartile range 277 – 1,608 days). | | |
| --- | --- | --- |
|  | Unadjusted | Adjusted for age, sex, clinical and exercise variables^†^ |
| PVC-/ECHO- | 1.0 | 1.0 |
| PVC+/ECHO- | 1.6 [0.8–3.2] | 1.2 [0.6–2.3] |
| PVC-/ECHO+ | 3.6 [2.1–6.0] | 1.7 [1.0–2.8] |
| PVC+/ECHO+ | 6.7 [4.1–10.9] | 2.3 [1.3–3.8] |
| Abbreviations: ECHO-: no significant abnormality on echocardiography; ECHO+: significant abnormality on echocardiography; PVC-: <1 PVCs/min during recovery; PVC+:≥1 PVCs/min during recovery.  ^†^Hypertension, diabetes, ischemic heart disease, heart failure, body-mass index, peak workload, maximal heart rate, heart rate recovery, ST depression, use of either betablocker, ace inhibitor, angiotension blockers, loop diuretics, antihrombotics, anti-coagulants, or calcium channel blockers. | | |

| **Table E. Sensitivity analysis – extended stratification based on PVC frequency**  Hazard ratios with 95% confidence intervals from Cox regression analysis, for the combination of PVCs during recovery (<1, ≥1–5/min, ≥5–10/min or >10/min) and echocardiographic abnormalities (n= 3,106, 219 events). | | |
| --- | --- | --- |
|  | Unadjusted | Adjusted for age, sex, clinical⸶ and exercise variables |
| PVC-/ECHO- | 1.0 | 1.0 |
| PVC-/ECHO+ | 4.3 [2.7–6.8] | 2.1 [1.3–3.3] |
| PVC >1–5/min/ECHO- | 2.6 [1.5–4.3] | 1.7 [1.0–2.9] |
| PVC >1–5/min/ECHO+ | 7.8 [5.0–12.1] | 3.3 [2.1–5.2] |
| PVC ≥5–10/min/ECHO- | 2.8 [1.0–8.0] | 1.6 [0.5–4.5] |
| PVC ≥5–10/min/ECHO+ | 10.9 [5.8–20.6] | 3.4 [1.7–6.6] |
| PVC >10/min /ECHO- | 1.2 [0.2–8.6] | 0.4 [0.1–3.1] |
| PVC >10/min /ECHO+ | 13.5 [6.7–27.3] | 3.7 [1.7–7.7] |
| Abbreviations: ECHO-: no significant abnormality on echocardiography; ECHO+: significant abnormality on echocardiography; PVC: premature ventricular contractions.  ^†^Hypertension, diabetes, ischemic heart disease, heart failure, body-mass index, peak workload, maximal heart rate, heart rate recovery, ST depression, use of either betablocker, ace inhibitor, angiotension blockers, loop diuretics, antihrombotics, anti-coagulants, or calcium channel blockers. | | |

| **Table F. Sensitivity analysis – echocardiography performed after or before the year 2010**  Hazard ratios with 95% confidence intervals from Cox regression analysis, for the combination of PVCs during recovery and echocardiographic abnormalities in who performed an echocardiographic study before (n=1,275) or after (n=2,346) Jan 2010. | | |
| --- | --- | --- |
| **Before January 2010** | | |
|  | Unadjusted | Adjusted for age, sex, clinical and exercise variables^†^ |
| PVC-/ECHO- | 1.0 | 1.0 |
| PVC+/ECHO- | 2.8 [1.3–6.0] | 1.6 [0.7–3.5] |
| PVC-/ECHO+ | 4.2 [1.3–6.0] | 1.8 [1.0–3.5] |
| PVC+/ECHO+ | 7.5 [4.1–13.8] | 2.7 [1.4–5.0] |
| **After January 2010** | | |
|  | Unadjusted | Adjusted for age, sex, clinical and exercise variables^†^ |
| PVC-/ECHO- | 1.0 | 1.0 |
| PVC+/ECHO- | 2.0 [0.8–4.7] | 1.2 [0.5–2.7] |
| PVC-/ECHO+ | 3.8 [1.8–7.8] | 1.6 [0.8–3.4] |
| PVC+/ECHO+ | 8.3 [4.2–16.1] | 2.8 [1.4–5.5] |
| Abbreviations: ECHO-: no significant abnormality on echocardiography; ECHO+: significant abnormality on echocardiography; PVC-: <1 PVCs/min during recovery; PVC+:≥1 PVCs/min during recovery.  ^†^Hypertension, diabetes, ischemic heart disease, heart failure, body-mass index, peak workload, maximal heart rate, heart rate recovery, ST depression, use of either betablocker, ace inhibitor, angiotension blockers, loop diuretics, antihrombotics, anti-coagulants, or calcium channel blockers. | | |

| **Table G. All-cause mortality**  Hazard ratios with 95% confidence intervals from Cox regression analysis, for the combination of PVCs during recovery and echocardiographic abnormalities in the prediction of all-cause death (n= 3,106, 506 events). | | |
| --- | --- | --- |
|  | Unadjusted | Adjusted for age, sex, clinical^†^ and exercise variables |
| PVC-/ECHO- | 1.0 | 1.0 |
| PVC/+/ECHO- | 2.1 [1.4–3.2] | 1.2 [0.9–1.7] |
| PVC-/ECHO+ | 2.7 [1.9–3.8] | 1.3 [0.9–1.7] |
| PVC+/ECHO+ | 3.6 [2.6–5.1] | 1.5 [1.1–2.0] |
| Abbreviations: ECHO-: no significant abnormality on echocardiography; ECHO+: significant abnormality on echocardiography; PVC-: <1 PVCs/min during recovery; PVC+: ≥1 PVCs/min during recovery  ^†^Hypertension, diabetes, ischemic heart disease, heart failure, body-mass index, peak workload, maximal heart rate, heart rate recovery, ST depression, use of either betablocker, ace inhibitor, angiotension blockers, loop diuretics, antihrombotics, anti-coagulants, or calcium channel blockers. | | |

| **Supplemental Table H**. Baseline characteristics and exercise stress test characteristics in patients who fulfilled inclusion criteria (no atrial fibrillation, exercise duration ≥3 min) but did not perform an echocardiographic examination | |
| --- | --- |
| Number of patients | 7,392 |
| Age, years | 56.7±14.0 |
| Male sex, n (%) | 3,981 (53.9) |
| Hypertension, n (%) | 711 (9.6) |
| Diabetes, n (%) | 415 (5.6) |
| Ischemic heart disease, n (%) | 558 (7.5) |
| of which, myocardial infarction, n (%) | 183 (2.5) |
| Cerebrovascular disease, n (%) | 79 (1.1) |
| COPD, n (%) | 137 (1.9) |
| Heart failure, n (%) | 33 (0.4) |
| *Medications* |  |
| ACE inhibitor, n (%) | 871 (11.8) |
| Betablocker, n (%) | 1400 (11.8) |
| Loop diuretics, n (%) | 132 (1.8) |
| Calcium antagonists, n (%) | 582 (7.9) |
| Thiazide diuretics, n (%) | 92 (5.2) |
| Anti-thrombotic, n (%) | 1,150 (15.6) |
| Nitrates, n (%) | 891 (12.1) |
| Anti-coagulant, n (%) | 64 (0.9) |
| *Exercise stress test variables* | |
| Peak workload, W | 169±61 |
| Peak workload, % of predicted | 92±18 |
| PVCs during recovery, n (%) |  |
| <1/min | 5,074 (68.6) |
| 1 – 5/min | 2086 (28.2) |
| 5 – 10/min | 155 (2.1) |
| >10/min | 77 (1.0) |
| PVCs during exercise, n (%) |  |
| <1/min | 4,694 (63.5) |
| 1 – 5/min | 2,297 (31.1) |
| 5 – 10/min | 246 (3.3) |
| >10/min | 155 (2.1) |
| Test terminated due to arrhythmia, n (%) | 27 (0.4) |
| Resting heart rate, beats/min | 75±13 |
| Maximum heart rate, beats/min | 155±23 |
| Heart rate recovery, beats/min | 32±14 |
| Maximum SBP, mm Hg | 195±26 |
| RPE (6 – 20), units | 17.3±1.1 |
| ST depression, n (%) | 345 (4.7) |
| Data presented as mean±standard deviation or n (%).  Abbreviations: ACE: angiotensin converting enzyme; COPD: chronic obstructive pulmonary disease; HF: heart failure; PVC: premature ventricular contractions; SBP, systolic blood pressure; W: Watts | |

**Supplemental Table I** –Referral questions^*^ (exercise stress testing)

| *Referral question* | *N (%)* |
| --- | --- |

| Coronary heart disease | 2354 (75.8) |
| --- | --- |
| Arrythmia | 279 (9) |
| Exercise tolerance | 212 (6.8) |
| Valvular heart disease | 131 (4.2) |
| Preoperative | 91 (2.9) |
| Dyspea | 44 (1.4) |
| Malignancy | 8 (0.3) |
| Control | 6 (0.2) |
| Transplantation | 3 (0.1) |
| Cardiomyopathy | 1 (1) |
| ECG findings | 1 (0) |
| Myocarditis | 1 (0) |
| Left ventricular function | 1 (0) |
| Syncope | 1 (1) |
| Other/missing | 471 (15.2) |
| ^*^Of note, referral question according to the clinical report does not equal diagnosis | |

| Supplemental Table J – Referral questions (echocardiography) | |
| --- | --- |
| *Referral question* | *N (%)* |
| LV function | 2492 (80.2) |
| Valvular disease | 266 (8.6) |
| Control | 198 (6.4) |
| None | 61 (0) |
| Peri-/myocarditis | 18 (0) |
| Cardiac embolism | 17 (0.5) |
| Pericarditis/pericardial effusion | 15 (0.5) |
| Endocarditis | 6 (0.2) |
| Heart murmur | 6 (0.2) |
| Dyspnea | 5 (0.2) |
| Cardiotoxicity | 5 (0.2) |
| Arrythmia | 4 (0.1) |
| Preoperative, valvular function | 3 (0.1) |
| Syncope | 3 (0.1) |
| Preoperative | 3 (0.1) |
| Exercise intolerance | 2 (0.1) |
| Pulmonary hypertension | 2 (0.1) |
| Vertigo/dizziness | 2 (0.1) |
| Transplantation | 2 (0.1) |
| Unspecific symptoms | 2 (0.1) |
| Aortaanerysm | 1 (0) |
| Heart failure | 1 (0) |
| Hypertension | 1 (0) |
| Myocarditis | 1 (0) |
| Palpitations | 1 (0) |
| Prior to coronary angiography | 1 (0) |
| Cardiomyopathy | 1 (0) |
| Coronary heart disease | 1 (0.0) |
| Other | 1 (0) |
| ^*^Of note, referral question according to the clinical report does not equal diagnosis | |
